# Combined RT-qPCR and Pyrosequencing of a SARS-CoV-2 Spike Glycoprotein Polybasic Cleavage Motif Uncovers Rare Pediatric COVID-19 Spectrum Diseases of Unusual Presentation

**DOI:** 10.1101/2020.12.19.20243428

**Authors:** Patrick Philipp Weil, Jacqueline Hentschel, Frank Schult, Anton Pembaur, Beniam Ghebremedhin, Olivier Mboma, Andreas Heusch, Anna-Christin Reuter, Daniel Müller, Stefan Wirth, Malik Aydin, Andreas C. W. Jenke, Jan Postberg

## Abstract

**Background:** Surveillance of severe acute respiratory syndrome coronavirus 2 (SARS-CoV-2) infections is essential for the global containment measures with regard to the ongoing pandemic. Diagnostic gold standard is currently reverse transcription of the (+)RNA genome and subgenomic RNAs and subsequent quantitative polymerase chain reaction (RT-qPCR) from nasopharyngeal swabs or bronchoalveolar lavages. In order to further improve the diagnostic accuracy, particularly for the reliable discrimination between negative and false-negative specimens, we propose the combination of the RT-qPCR workflow with subsequent pyrosequencing of a S-gene amplicon. This extension might add important value mainly in cases with low SARS-CoV-2 load, where RT-qPCR alone can deliver conflicting results.s

**Results:** We successfully established a combined RT-qPCR and S-gene pyrosequencing method which can be optionally exploited after routine diagnostics or for epidemiologic studies. This allows a more reliable interpretation of conflicting RT-qPCR results in specimens with relatively low viral loads and close to the detection limits of qPCR (C_T_ values >30). After laboratory implementation and characterization of a best practice protocol we tested the combined method in a large pediatric cohort from two German medical centers (n=769). Pyrosequencing after RT-qPCR enabled us to uncover 6 previously unrecognized cases of pediatric SARS-CoV-2 associated diseases, partially exhibiting unusual and heterogeneous presentation. Moreover, it is notable that in the course of RT-qPCR/pyrosequencing method establishment we did not observe any case of false-positive diagnosis when confirmed SARS-CoV-2-positive specimens were used from foregoing routine testing.

**Conclusions:** The proposed protocol allows a specific and sensitive detection of SARS-CoV-2 close to the detection limits of RT-qPCR. Combined RT-qPCR/pyrosequencing does not negatively affect preceding RT-qPCR pipeline in SARS-CoV-2 diagnostics and can be optionally applied in routine to inspect conflicting RT-qPCR results.

## Background

As of April 26^th^ 2020 at Helios University Hospital Wuppertal during a period of attenuation which followed the first peak phase of SARS-CoV-2 transmission in Germany only 2.3% of all laboratory-confirmed SARS-CoV-2-positive cases were children or teenagers (age group: 0-19 yrs). Nationwide as of September 8^th^ 2020, the contribution of this age group to all SARS-CoV-2-positive cases had increased to 10.5% [1]. This development was in agreement with observations made globally, in particular in the US, whose population remains among the most affected of the SARS-CoV-2 pandemic. Since then, a trend of weekly median age decline for persons with COVID-19-like illness was reported for an observation period between May 3^rd^ – August 29^th^ 2020. In particular, a steady increase of the percentage fraction of all confirmed SARS-CoV-2-positive cases for the 0-19 year olds was observed in the United States starting from 7.4% in May, 10.8% in June, 14.0% in July and preliminary being elevated up to 15.5% in August 2020 [2]. Reminiscent of this, at least since early autumn 2020 the 7-days-incidence for all pediatric age groups increased in similar ways as observed for most other age groups in Germany (Figure 1). As evidence is accumulating that pediatric presentation of SARS-CoV-2 associated diseases can possibly be more heterogeneous than adult COVID-19 there is an urgent need to recognize the full spectrum of unusual pediatric SARS-CoV-2-borne diseases. Reliable diagnosis of SARS-CoV-2 infection will eventually contribute to effective personalized treatment and optimized containment measures. To address this problem, we systematically screened two pediatric cohorts from Helios University Hospital Wuppertal (North Rhine-Westphalia, Western Germany) and Klinikum Kassel (Hessen, Central Germany). Therefore, in order to enable the re-examination of ambiguous SARS-CoV-2 (+)RNA RT-qPCR results, which particularly can occur in specimens with relatively low viral load, we developed and applied an improved assay that combines RT-qPCR and pyrosequencing. This strategy allows the reliable discrimination of SARS-CoV-2 from other human coronaviruses (HCoVs) and can reduce the rate of uncertain qPCR results, which might be influenced by several confounding factors, thus occasionally leading to false-negative qPCR results – particularly when PCR is close to its detection limits. Here, we provide a preliminary report of our epidemiological survey as well as a detailed protocol for an expanded SARS-CoV-2 (+)RNA detection method, which relies on the complementary exploitation of RT-qPCR and pyrosequencing of a genome fragment from the SARS-CoV-2 Spike glycoprotein polybasic cleavage motif. Following rigorous implementation, we applied the test on two pediatric cohorts from two German medical centers comprising 769 children in total (n=599 [Wuppertal]; n=170 [Kassel]; 44% female/56% male; 53.6% 0-5 yrs, 28.7% 6-12 yrs, 17.7% 13-17 yrs) without indication for routine COVID-19 testing as defined by the WHO at time of presentation.

**Figure 1.**
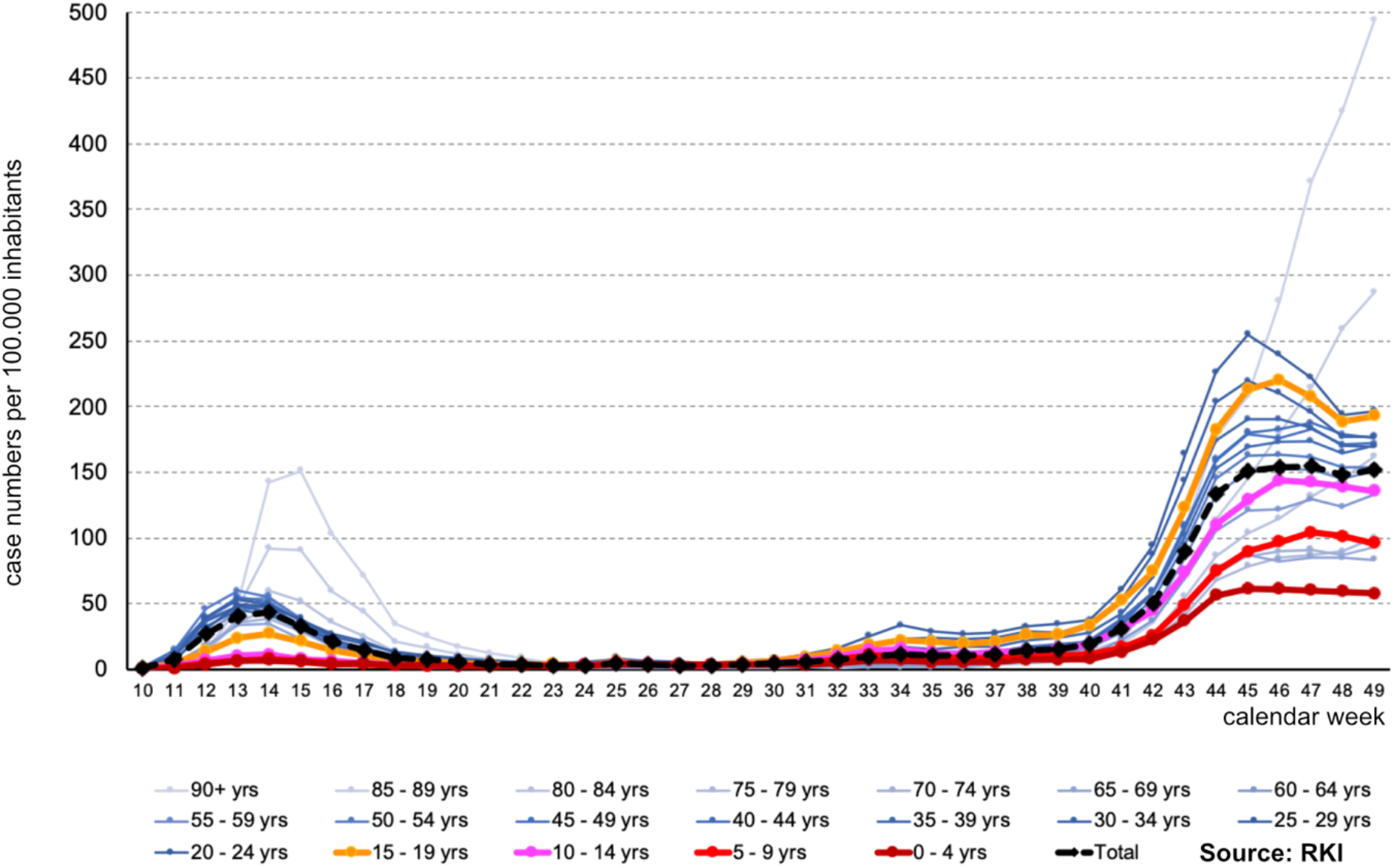
*7-days incidence development in Germany between 2020 calendar weeks 10-49*. According to the laboratory confirmed SARS-CoV-2 case numbers continuously reported by the Robert-Koch-Institute (www.RKI.de) the 7-days-incidence for pediatric infections increased very reminiscent of most other age-groups. At least for the age groups between 0-14 years, this development contrasts the spring situation, when these children were less affected than other age-groups.

## Results

### Design and implementation of a combined SARS-CoV-2 RT-qPCR/pyrosequencing test to complement established diagnostic workflows

We targeted a region encoding a polybasic cleavage motif within the SARS-CoV-2 Spike glycoprotein (S). Multiple sequence alignment analyses demonstrate that this region could be a suitable differentiator between SARS-CoV-2, related coronaviruses from several animal host [3] as well as SARS-CoV-1, MERS-CoV and other HCoVs (Figure 2A.). For RT-qPCR we selected a set of forward primer (S_pbc_-CoV-2-F: 5’-GCAGGCTGTTTAATAGGGGC-3’) and 5’-biotinylated reverse primer (S_pbc_-CoV-2-R_BIO_: 5’-biotin-TEG-ACCAAGTGACATAGTGTAGGCA-3’), since Primer-BLAST (https://www.ncbi.nlm.nih.gov/tools/primer-blast/index.cgi) query confirmed the probable suitability of the selected primer set predicting a 162 bp SARS-CoV-2-specific amplicon matching perfectly to the complete reported list of SARS-CoV-2 isolates. As probe for TaqMan qPCR we used 5’-HEX-ATTGGTGCAGGTATATGCGCTAGTTATC-BBQ-650-3’ (S_pbc_-CoV-2-P). Without carrying the 5’-/3’-modifications we selected the same oligonucleotide sequence for the pyrosequencing primer (S_pbc_-CoV-2-S) (Figure 2B.; Table S1).

**Figure 2.**
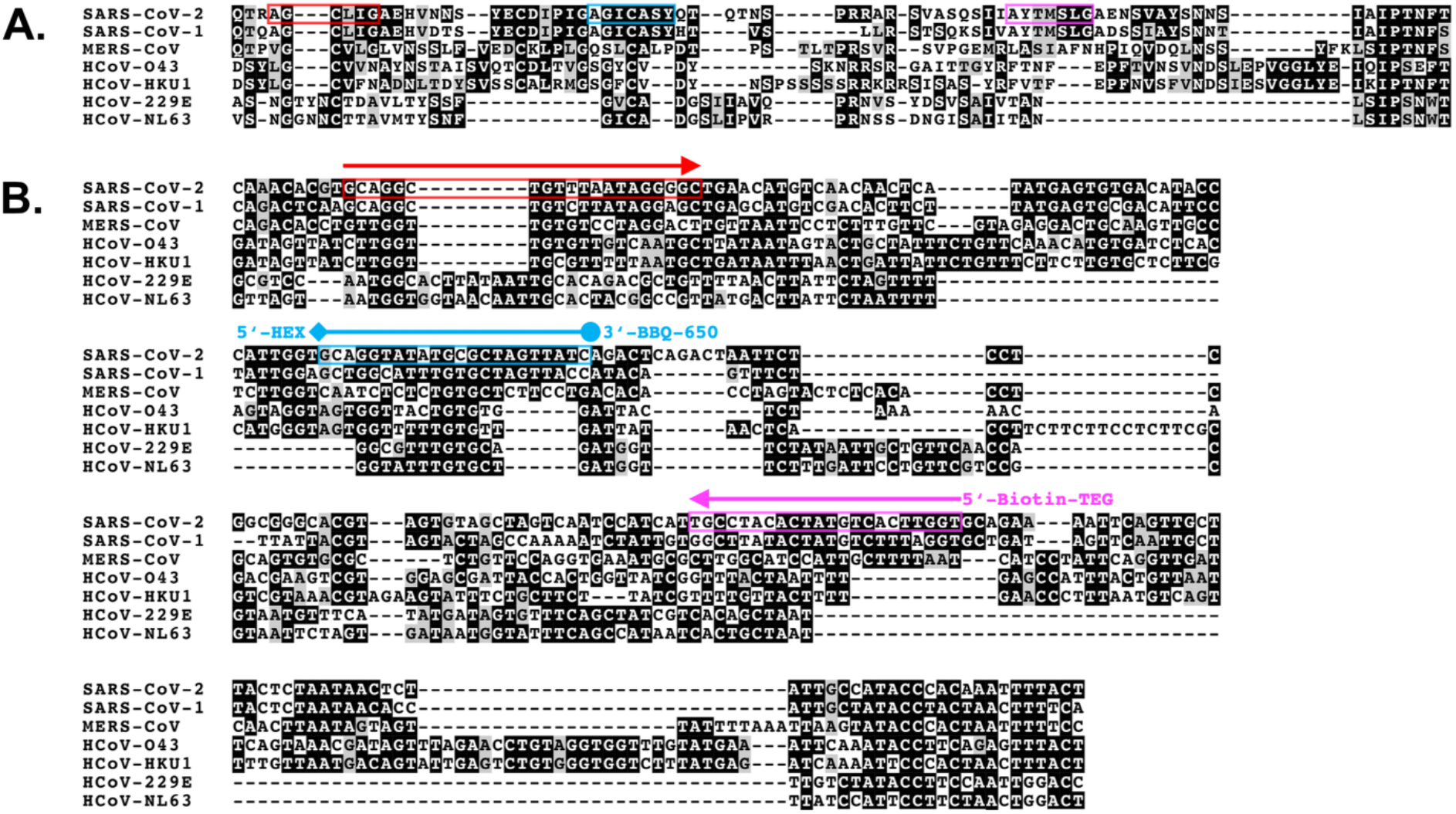
*Comparison of homologous protein sequence segments* **(A**.) *and corresponding cDNA segments* **(B**.) *between human coronaviruses*. **A**. We applied the Clustal W algorithm of MEGA [17] to conduct multiple-sequence alignments using the translated protein sequences of the SARS-CoV-2 Spike (S) glycoprotein and homologous S protein sequences from other human coronaviruses. Here, a segment harboring polybasic cleavage motif (Q644 to T720 with respect to SARS-CoV-2 S protein). The highlighted residues are conserved in most human coronavirus (black shaded) or are similar between some human coronaviruses (grey shaded). The colored boxes are framing residues, which correspond to the target position of tested oligonucleotides: forward primer (red), probe/sequencing primer (blue), reverse primer (magenta). **B**. The aligned protein sequences were backtranslated into the encoding cDNA sequences. Similarly, as described above for protein sequences, black or grey shading was used to illustrate identical or similar nucleotide positions. For combined RT-qPCR and pyrosequencing, we selected the following marked sequences for oligonucleotide design: 1. Forward primer (red box/arrow); 2. TaqMan probe with 5’-HEX and 3’-BBQ-650 modifications (blue box/line); 3. Sequencing primer without end-modification (blue box – same sequence as TaqMan Probe); 4. Reverse primer with 5’-Biotin-TEG (magenta box/arrow).

For all tests we used specimens, which underwent routine COVID-19 diagnostic testing and were pre-categorized ‘confirmed SARS-CoV-2-positive’ or ‘negative’. For comparison we used the full set of ‘Charité protocol’ amplicon targets (RdRP, E, N) [4] (Table S1). Before RT-qPCR, total RNA was purified from 250 µL of nasopharyngeal swabs or bronchoalveolar lavage specimens using the guanidinium isothiocyanate (GITC) extraction method. Besides, due to RT-qPCR reagent manufacturer’s recommendation, we tested whether the RNA purification step could be dispensable. Briefly, using stabilized raw specimens from SARS-CoV-2 as RT-qPCR templates, results from the same specimens using purified RNA could only be matched in some cases (Figure S1). Therefore, we neglected this approach during our successive experiments.

Reverse transcription and subsequent amplification of cDNA were carried out using universal one-step RT-qPCR reaction mastermixes. A detailed protocol including recommended reagents is provided in the Methods section. Quantitative PCR programs often use an excess of cycling steps, frequently leading to the incremental enrichment of unspecific byproducts during later cycles. With respect to the quality of subsequent pyrosequencing, we aimed on the optimization of PCR cycle numbers for singleplex, duoplex and triplex PCR approaches. We determined that 36 sequential qPCR cycles of denaturation, annealing and elongation were a good compromise with respect to multiple prudential reason: 1. Typically, for different amplicons we observed that the cycle threshold (C_T_) limit for the faithful discrimination between positive and negative samples was below a C_T_ of 35 (Figure 3).

**Figure 3.**
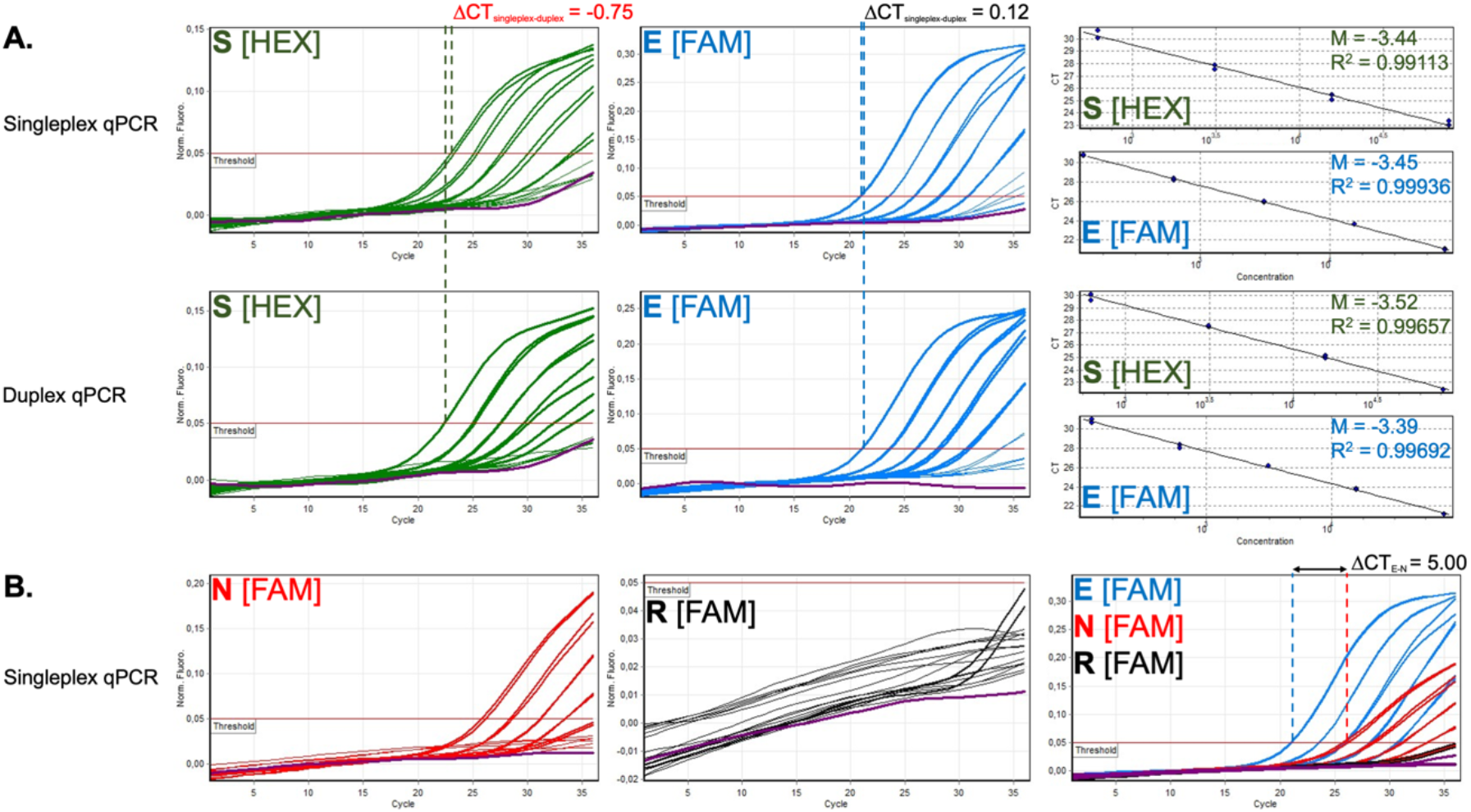
Comparative characterization of RT-qPCR efficiency and sensitivity. The same clinical specimen was used for all tests. **A**. The S-gene amplicon (S) was compared with ORF E (E) in singleplex and duoplex reactions, respectively. **B**. The direct performance comparison of ORF E (E) amplicons with ORF N (N) and RdRP (R) demonstrated the differences in sensitivity, whereby E>N>R.

Consistently, semi-quantitative PCR and subsequent electrophoresis resulted in a specific 162 bp band and no or occasionally few weak byproducts (Figure S2A). 2. For approx. 35 or more cycles we observed accelerated curve increments and thresholds crossing also for negative samples and no-template controls indicating ongoing amplification of PCR byproducts. Using an excess of PCR cycles, multiple unspecific byproducts and DNA smear of higher molecular weight are observable in confirmed positive and negative specimens (Figure S2B).

Besides an excess of PCR cycles, we identified multiplex PCR approaches where more than one amplicon is targeted by multiple primer sets in one PCR reaction as another possible confounding factor influencing PCR quality and successive pyrosequencing. To test this hypothesis, we performed triplex RT-qPCR reactions by simultaneously using RdRP, Orf E and Orf N amplicon targets. Semi-quantitative analyses by agarose gel electrophoresis demonstrate that numerous low and high molecular weight byproducts were amplified in both cases when RNA from confirmed SARS-CoV-2-positive specimens or negative specimens was used (Figure S3). Since we assumed that biotinylated byproducts could massively impair successive pyrosequencing we decided to neglect triplex approaches for the intended combinatorial use of RT-qPCR. With emphasis on the SARS-CoV-2 S-gene amplicon we instead aimed to compare the specificities, efficiencies and sensitivities of singleplex RT-qPCR vs. duoplex RT-qPCR. In order to further characterize the performance of the SARS-CoV-2 S-gene amplicon target and to select the best additional amplicon for a duoplex approach, we performed comprehensive RT-qPCR tests using the S-gene, Orf E, Orf N and RdRP as amplicon targets and using serially diluted specimens from the same confirmed SARS-CoV-2-positive specimens for all RT-qPCR reactions. Figure 3 illustrates the performance of different qPCR assays for one representative SARS-CoV-2-positive nasopharyngeal specimen. Taken together, TaqMan RT-qPCR for both the S-gene amplicon and the Orf E amplicon with similar sensitivity as indicated by low ΔC_T_ between S-gene and Orf E in singleplex reactions. Further, we deduced from a series of serial template dilutions that amplification efficiency for both the S-gene target and the Orf E target is high but weakens between C_T_=30 and C_T_=35. In duoplex RT-qPCR assays targeting simultaneously the S-gene target and Orf E in one reaction no notable changes in sensitivity or qPCR efficiency were seen (Figure 3A). With respect to efficiency also the Orf N amplicon target performed well, but its sensitivity lagged behind as indicated by a large ΔC_T_ when compared to Orf E (Figure 3B). In stark contrast, the RdRP amplicon frequently performed volatile and appeared to lag behind the sensitivities of all other amplicons in the majority of specimens examines - as in the illustrated case, where even the curve for the undiluted sample did not cross the predefined threshold (Figure 3B). Therefore, apart from a singleplex S-gene RT-qPCR/pyrosequencing approach, we considered a duoplex (S-gene and Orf E) RT-qPCR/S-gene pyrosequencing approach as a promising combination for the faithful detection of active SARS-CoV-2 infections.

To test this hypothesis, we performed subsequent pyrosequencing of the biotinylated single-stranded S-gene amplicon using the same serial dilution samples described above (Figure 4). The proposed pyrosequencing approach could theoretically allow to read a SARS-CoV-2 (+)RNA-specific sequence fragment of 55 nt. Using the undiluted sample from the singleplex S-gene amplicon RT-qPCR reaction, we achieved an unbiased 53 nt sequence fragment, which could unambiguously be assigned to the SARS-CoV-2 reference genome. Successive serial dilutions led to modest enrichment of miscalled bases in the resulting pyrogram, but still allowed the faithful assignment of the called sequence to SARS-CoV-2 up to a corresponding approx. C_T_≈30. Serial dilutions were gradually accompanied by a weakening of pyrogram-signal strength and a decreased signal-to-noise ratio. Automated basecalling of the SARS-CoV-2 sequence failed frequently for samples with C_T_>30. However, for a range between C_T_≈30 and C_T_≈35 the most adverse reason counteracting SARS-CoV-2 sequence recognition appeared to be the decremental signal-to-noise ratio, which led to incremental false-positive calling of repetitive bases. The illustrated examples suggest the presence of signals encoding the targeted SARS-CoV-2, which could be separable from the threshold noise signals. S-gene pyrosequencing performed equally well, when a duoplex (S-gene and Orf E) RT-qPCR was conducted priorly (Figure 4). We noted a tendency of slightly increasing numbers of missing or excess basecalls towards the 3’-end of the pyrosequenced SARS-CoV-2 fragment. As a preliminary conclusion we first highlight that the proposed pyrosequencing approach does not negatively affect preceding RT-qPCR pipeline in SARS-CoV-2 diagnostics. Second, it adds important value to RT-qPCR, where this method alone delivers conflicting results, particularly close to the detection limits qPCR (C_T_ values >30). In RT-qPCR alone even negative samples frequently exhibit curves crossing the threshold within a range between C_T_≈30 and C_T_≈35 complicating mainly the reliable discrimination between PCR-negatives and PCR-false-negatives.

**Figure 4.**
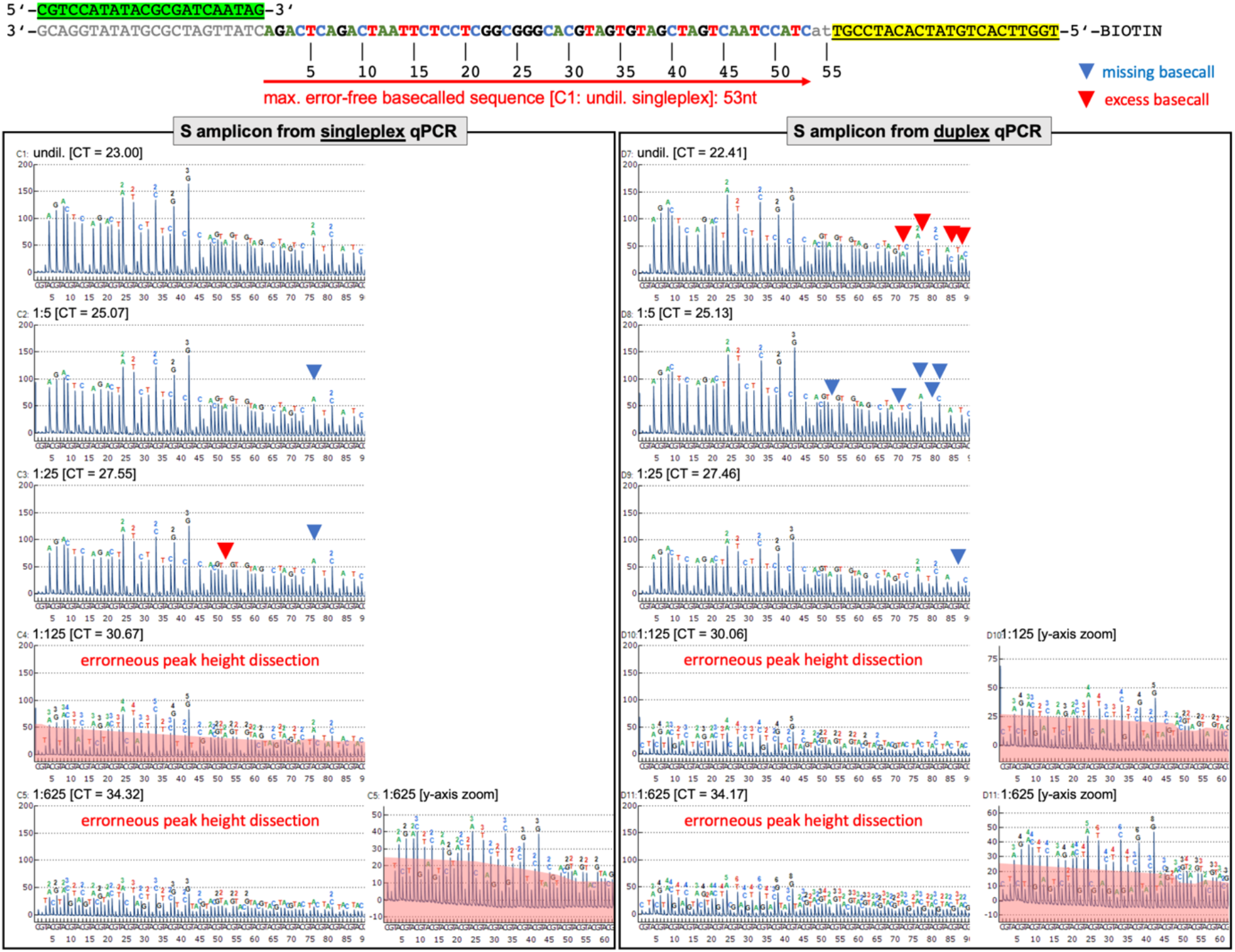
Comparative characterization of pyrosequencing sensitivity and basecalling quality. Serial dilutions of the same clinical specimen were used for all tests. **Top**. Targeted region and principle of S-gene fragment pyrosequencing. Limited by the used PCR primers the maximum theoretical sequence length is 55 nt. **Bottom left**. Resulting pyrograms from serial template dilutions used for RT-qPCR in singleplex reactions and subsequent pyrosequencing of the S-gene amplicons using the antisense single-strand (as defined by the biotinylated reverse primer [S_pbc_-CoV-2-R_BIO_]) are shown. From the undiluted singleplex reaction we obtained the longest unbiased SARS-CoV-2-specific sequence fragment, which had an error-free length of 53 nt. **Bottom right**. Resulting pyrograms from serial template dilutions used for RT-qPCR in duoplex reactions and subsequent pyrosequencing of the S-gene amplicons using the antisense single-strand (as defined by the biotinylated reverse primer [S_pbc_-CoV-2-R_BIO_]) are shown. From the undiluted duoplex reaction we obtained a maximum unbiased SARS-CoV-2-specific sequence fragment of 44 nt in length. **Bottom left/right**. The associated C_T_ values are shown besides the degree of dilution. Lower template concentrations led to the occasional occurrence of miscalled bases (missing bases: red triangle; excess bases: blue triangle) and gradual convergence of signal and noise peaks. Whereas concomitantly, automated basecalling gradually failed to separate signal from noise, the SARS-CoV-2 specific sequence could be identified by manual inspection much longer.

In a next step, after protocol implementation we checked the sustainability of the combined RT-qPCR/pyrosequencing method in an epidemiological field test.

### Performance of the combined methods of RT-qPCR and pyrosequencing in epidemiologic field study on pediatric patients from two German medical centers

We tested our combined RT-qPCR/pyrosequencing method on two pediatric cohorts from two German medical centers comprising 769 children in total (n=599 [Wuppertal]; n=170 [Kassel], who had no indication for routine SARS-CoV-2 testing at presentation according to WHO criteria. After initial S-gene amplicon detection, we used at least ORF E as backup amplicon as well as ORF N and RdRP in some cases. Even for RT-qPCR results with relatively high C_T_ values, successive pyrosequencing could unambiguously confirm SARS-CoV-2 infections. Where possible, we investigated SARS-CoV-2-specific immunity. In sum, using retrospectively the combined RT-qPCR/pyrosequencing method, we confirmed 6 pediatric cases of SARS-CoV-2-associated diseases among the entire cohort exhibiting heterogenous presentation:

### *Brief case report 1* Adolescent male with sore throat (age group 13-17 years)

During the 2020 spring peak of incremental SARS-CoV-2 transmission at end of April an adolescent male patient presented at the emergency department in slightly reduced general condition with 37.5°C body temperature and reddened throat. The clinical examination was otherwise completely normal. At time of presentation the patient did not fulfill the criteria for routine SARS-CoV-2 testing. The specimen was incidentally tested and classified potentially being SARS-CoV-2-positive in the course of our serial examination of the Wuppertal pediatric cohort using the newly developed S-gene amplicon target for singleplex RT-qPCR (repetitive C_T_ values: 25.47 and 25.33). The result was unambiguously confirmed by pyrosequencing and backed-up by positive RT-qPCR results using the ORF E (C_T_ value: 23.78), ORF N (C_T_ value: 27.47) and RdRP (C_T_ value: 31.23) amplicon targets. Three months after first presentation, the patient was positive for SARS-CoV-2-specific antibodies (34.20 +; Roche), strongly suggesting that the boy underwent an inconspicuous course of SARS-CoV-2 infection.

### *Brief case report 2*. Female toddler with Multisystem Inflammatory Syndrome in Children (MIS-C) (age group 1-3 years)

A female toddler was admitted to the hospital with altered general status and undulant fever. The initial physical examination revealed tonsillitis without any cardio-respiratory affections. Laboratory analyses revealed highly elevated C reactive protein (24.0 mg/dL [normal<0.5]) with almost normal interleukin-6 levels at 55.2 pg/ml. Interestingly, no leukocytosis or lymphopenia were diagnosed. In the clinical course, symptoms reminiscent of Kawasaki-like disease included persistent fever, bilateral conjunctivitis, cheilitis and a maculopapular exanthema. Furthermore, echocardiography exhibited enlargement of the left coronary artery and pericardial effusion (Figure S4A). Cardiac related blood parameters were within the normal ranges. Under the suspicion of Multisystem Inflammatory Syndrome in Children (MIS-C) associated with COVID-19 [5, 6] she was given intravenous gamma globulins (2 g/kg), prednisolone (2 mg/kg) and acetylsalicylic acid (50 mg/kg) at day five which resulted in rapid improvement of the girl’s general status. Apyrexia was achieved on day seven. At day ten of hospitalization SARS-CoV-2 RT-qPCR tests were negative.

### *Brief case report 3*. Pre-school boy with high grade fever (age group 4-6 years)

A pre-school boy with bowel disease history presented at the paediatric emergency department with a history of one day high-grade fever. Clinical examination did not reveal a specific focus. He was admitted for suspected sepsis and started on intravenous antibiotics. After one day the condition of the patient and the inflammatory markers improved. The blood culture confirmed a bacterial bloodstream infection (BSI). On day 2 of inpatient care, the patient developed facial, neck and upper limb oedema, vein distention in the upper chest and shortness of breath suggestive of superior vena cava (SVC) syndrome. CT-angiography of the chest confirmed this diagnosis demonstrating a stenosis of the SVC (lumen diameter 1.4 mm), adjacent thrombosis and multiple collateral veins draining towards an enlarged vena azygos. Retrospectively, we uncovered a co-infection with SARS-CoV-2, which presumably was temporally associated with the BSI. Mean C_T_ value of repetitive RT-qPCR was relatively high (C_T_: 34.06) with conspicuous curve shape. SARS-CoV-2 was unambiguously confirmed by pyrosequencing.

### *Brief case report 4*. Male secondary school child with isolated swollen neck lymph node (age group 10-12 years)

A male prepubescent secondary school child presented to the emergency department in late March 2020 complaining about a ‘bump’ behind his left ear. Otherwise, he was completely asymptomatic. Clinical examination revealed a non-tender swelling behind the left ear of about 7-8 mm in diameter which was interpreted as an enlarged lymph node as well as slightly enlarged cervical lymph nodes on the same site. No cardiac, neurological, pulmonary or gastrointestinal abnormalities were note and the patient was sent home. At time of presentation the patient did not fulfill the criteria for routine SARS-CoV-2 testing. Mean C_T_ value of repetitive RT-qPCR was relatively high (C_T_: 35.56) with conspicuous curve shape. SARS-CoV-2 was unambiguously confirmed by pyrosequencing.

### *Brief case report 5*.: Female adolescent with EBV-like disease (age group 14-17 years)

A female adolescent presented late in March 2020 with high fever up to 40.2°C, hepatomegaly and tonsillitis. She was found to have acute Epstein-Barr-Virus infection and was discharged home 3 days later without fever in good general condition. Nine days later she presented again with low temperature up 38.4°C, general malaise and slightly increased liver enzymes. All other diagnostic test at that time – including abdominal ultrasound, laboratory studies and urine analysis – were normal. During the following two days she improved spontaneously and was discharged home. At that time symptoms were believed to be associated with the recent EBV infection. No SARS-CoV-2 testing was performed since the patient did not fulfill the clinical criteria for suspected Covid-19 at that time. Mean C_T_ value of repetitive RT-qPCR was relatively high (C_T_: 32.80) with conspicuous curve shape. SARS-CoV-2 was unambiguously confirmed by pyrosequencing. In October 2020 the patient was found to have IgA and IgG antibodies against SARS-CoV-2 (SARS-CoV-2 ELISA, EUROIMMUN).

### *Brief case report 6:* Male toddler with generalized febrile seizure (age group 1-3 years)

A male toddler was admitted late in March 2020 due to a generalized febrile seizure which spontaneously resolved after 4 min. The parents reported that he developed a low-grade fever up to 38.6°C several hours prior the event. He also had moderate diarrhoea with three to four pulpy bowel movements per days and vomited three times on the day of admission. The parents did not note any mucous or blood. On admission, clinical examination was unremarkable except for mild pharyngitis, intestinal hyperperistalsis and mild dehydration. Differential blood count, liver enzymes, blood gases, electrolytes, C-reactive protein as well as urine analyses (except for ketone bodies) were all normal. Stool tests came back negative for Rotavirus, Norovirus, Shigella, Campylobacter, Salmonella and Yersinia. The patient received intravenous fluids and was discharged two days later without fever in good clinical condition. At that time the patient was diagnosed with mild gastroenteritis and secondary uncomplicated generalized febrile seizure. RT-qPCR and pyrosequencing confirmed a SARS-CoV-2 infection.

## Discussion

### Numerous confounding factors influence the outcome of RT-qPCR in SARS-CoV-2 diagnostics

Whereas reverse transcription and subsequent quantitative PCR (RT-qPCR) are key methods for the detection of SARS-CoV-2 and local as well as global pandemic surveillance, it is well known that several confounding factors can lead to false-negative results. It seems clear that the time course of SARS-CoV-2 load in the days after infection massively influences the predictive value of the RT-qPCR tests. A recent study discovered a median false-negative rate as high as 38% (CI, 18% to 65%) even at the day of symptom onset, although concomitantly SARS-CoV-2 load seems to be close to its highest levels. Moreover, few days before and after symptom onset the false-negative SARS-CoV-2 discovery rate by RT-qPCR seems to worsen dramatically [7]. Besides the time point of specimens sampling post-infection, heterogeneities in specimens sampling technique, transportation, storage conditions, nucleic acids purification, laboratory equipment, staff experience but also RT-qPCR conditions might be among the most important factors influencing test qualities. Very early in 2020 the WHO started to disseminate the ‘Charité protocol’ for the diagnostic detection of SARS-CoV-2 by RT-qPCR. Therein, a strategy for the combinatorial use of PCR primers (#name_F/#name_R) and TaqMan probes (#name_P) was described with amplicon targets in the SARS-CoV-2 RNA-dependent RNA polymerase (RdRP) gene as well as in the open reading frames (ORF) E and N (E_Sarbeco_F/_P1/_R; N_Sarbeco_F/_P/_R; RdRP_SARSr-F/_P2/_R) (Table S1) [8]. As of November 10^th^ 2020, PubMed reported 976 citations for this article, suggesting the widespread use of the contained protocols for SARS-CoV-2 detection in association with the global attempt of pandemic surveillance and containment measures. However, several RT-qPCR assays are being used by clinical, research and public health laboratories and some studies suggest that there could be relevant differences in the efficiencies, sensitivities and specificities of the different amplicon targets under different laboratory conditions, in particular for the RdRP_SARSr amplicon [9, 10]. Remarkably, RT-qPCR does not exclusively target genomic SARS-CoV-2 amplicons, but (dependent on the selected primers) logically also can amplify the transcribed shorter subgenomic mRNAs (sgRNAs). At least two deep-sequencing studies on coronavirus transcriptomes (HCoV-229E and SARS-CoV-2) reported a dominant coverage of mapped reads towards the 3’-end of the SARS-CoV-2 genome, possibly due to an abundance of the sgRNAs [11, 12]. Notably, these sgRNAs encode conserved structural proteins with the Spike glycoprotein (S), the envelope protein (E) and the nucleocapsid protein (N) being among them. For the nonstructural RdRP, in contrast, the genomic (+)RNA itself becomes expressed by genome translation and ribosomal frameshifting [13]. The coverage of mapped reads was comparatively lower for the RdRP encoding ORF1b as well as the 5’-proximal ORF1a than for the sgRNA encoding 3’-end. However, it remains poorly understood so far how oscillations in the expression of SARS-CoV-2 sgRNAs might contribute to the observed variations in qPCR testing.

Therefore, we aimed to develop a confirmatory test for a stably expressed SARS-CoV-2 sgRNA amplicon target that can complement RT-qPCR strategies without disturbing established and automatable laboratory workflows. In particular, we intended to develop a pyrosequencing assay that would allow, subsequent to RT-qPCR, the categorical confirmation of SARS-CoV-2 infections in acute cases, where the clinical suspicion is high, but the SARS-CoV-2 infection cannot be ruled out by RT-qPCR alone. The proposed pyrosequencing approach does not negatively affect preceding RT-qPCR pipelines in SARS-CoV-2 diagnostics and can therefore add important value to RT-qPCR, where this method alone delivers conflicting results. Particularly, this can happen close to the detection limits qPCR, practically C_T_ values >30. Frequently, even negative samples exhibit curves crossing the threshold within a range between C_T_≈30 and C_T_≈35 complicating the reliable discrimination between PCR-negative and PCR-false-negative specimens. Theoretically, also PCR-false-positives results could happen, but practically we did not observe any case of false-positive diagnosis within the numerous confirmed SARS-CoV-2-positive specimens from clinical routine testing used during the entire phase of RT-qPCR/pyrosequencing development. Here, we have shown that pyrosequencing can be a powerful complementary method of specific and sensitive SARS-CoV-2 case confirmation, without affecting foregoing routine RT-qPCR. But even here, lower template concentrations led to the occasional occurrence of miscalled bases (missing bases: red triangle; excess bases: blue triangle) and gradual convergence of signal and noise peaks. Gradually, this led to increasingly misinterpreted peak heights particularly for nucleotide repeat motifs and lower complexity motifs. Whereas concomitantly, automated basecalling gradually failed to separate signal from noise, the SARS-CoV-2 specific sequence could be identified by manual inspection much longer. Manufacturer’s adaptations to the basecalling algorithm could lead to an improvement. Moreover, for each nucleotide position we used an ‘open’ pyrosequencing dispensation order (assuming N [i.e. C, G, A or T] as possible nucleotides for each position) in order to detect variable positions. From the end users’ sides, a SARS-CoV-2 specific dispensation order could possibly lead to an improved automated basecalling sensitivity. However, being aware that any further sample treatment in laboratory routine would impair the necessary high throughput testing, in particular in the course of SARS-CoV-2 surveillance, we see the main application opportunities for combined RT-qPCR/pyrosequencing in research for retrospective epidemiology studies, longitudinal studies of the course of COVID-19 cases and studies on pathomechanisms, in particular if they aim on systemic surveys of virus-host interaction.

### Several cases with unusual presentations of pediatric SARS-CoV-2-infections were uncovered

Exemplarily for the implementation of the developed experimental pipeline for an epidemiological survey, we conducted a field study in search for SARS-CoV-2 infected patients without obvious common SARS-CoV-2 associated symptoms, since the majority of cases pediatric SARS-CoV-2 infections develop only very mild disease courses [14]. In a very recent UK national cohort study on neonatal SARS-CoV-2 infection 66 SARS-CoV-2-positive babies could be identified, who at day of presentation exhibited hyperthermia, poor feeding, vomiting, coryza, other respiratory signs and lethargy as the most common signs of infection or, respectively, no signs of infection at all [15]. On the other hand, an unknown fraction of SARS-CoV-2 infections in childhood seem to fundamentally differ from adults and can be more heterogeneous in their presentation. The most striking example is Multisystem Inflammatory Syndrome in Children (MIS-C), a rare SARS-CoV-2-induced Kawasaki-like hyperinflammatory syndrome [5, 6]. Another recent study reports that children and adults can exhibit a very different antibody responses upon SARS-CoV-2-infections across the clinical spectrum of associated diseases, which do not obligatorily match the adult COVID-19 spectrum [16].

Necessarily, the association unusual symptoms with acute infections will contribute to our understanding about the heterogeneity of SARS-CoV-2-borne diseases in general and particularly in children. Using two large pediatric cohorts for a field study, we have successfully demonstrated in this study that combined RT-qPCR/pyrosequencing is a reliable tool that allows the faithful confirmation of less distinctive or asymptomatic cases with SARS-CoV-2 infection close to the detection limits of RT-qPCR. Here, the determination of a SARS-CoV-2-specific sequence fragment can decisively help to improve reliability for cases where the discrimination between negative/false-negative RT-qPCR reports can be important. Whereas this might possibly be of subordinate importance for practical containment measures because low viral load could be associated with low infectiousness, it can contribute to recognize the still probably underestimated extent of SARS-CoV-2 prevalence and associated mild or asymptomatic presentations. In particular, from a large cohort of children who did not fulfil the criteria for SARS-CoV-2 testing we have retrospectively identified 6 cases of SARS-CoV-2 related symptoms. The identification and characterization of this cases will thus contribute to the understanding of the full heterogenous picture of presentation of SARS-CoV-2 infections.

## Conclusions

The proposed protocol allows the specific and sensitive detection of SARS-CoV-2 close to the detection limits of RT-qPCR. Combined RT-qPCR/pyrosequencing does not negatively affect preceding RT-qPCR pipeline in SARS-CoV-2 diagnostics and can be optionally applied in routine to inspect conflicting RT-qPCR results. Particularly in research, the proposed workflow can valuably contribute to identify and characterize the unknown fraction of cases of SARS-CoV-2 -associated diseases in childhood, which can fundamentally differ from adults and can be more heterogeneous in their presentation.

## Methods

### Sampling of pediatric and adult human specimens

Nasopharyngeal swabs or bronchoalveolar lavage specimens were collected at Helios University Hospital Wuppertal (North Rhine-Westphalia, Western Germany) and Klinikum Kassel (Hessen, Central Germany) with approval of the Witten/Herdecke University ethics commitee (covered by ‘CoronaKids’ [No. 61/2020] and No. 160/2020 for the use of routinely sampled and confirmed specimens for RT-qPCR/pyrosequencing method establishment). Informed written consent was obtained from legal guardians of the involved children. All work has been conducted according to the principles expressed in the Declaration of Helsinki.

### Storage and nucleic acids isolation

Specimens included nasopharyngeal swabs or bronchoalveolar lavage specimens, which underwent routine COVID-19 diagnostic testing. Pediatric cohort specimens were collected using brushes from the Gentra Puregene Buccal Cell Kit (100) (Qiagen, Cat. No. 158845) and then stabilized in 500 µL RNAlater™ (Thermo Fisher Scientific, Cat. No. AM7021). All specimens were stocked at -80°C. Total RNA was purified from 250 µL liquid specimen using 750 µL QIAzol lysis reagent (Qiagen, Cat. No. 158845) upon manufacturer’s recommendations. RNA quality and quantity were assessed by microcapillary electrophoresis using the Small RNA kit (Agilent, Cat. No. 5067-1548) and the Agilent Bioanalyzer 2100 instrument.

Importantly, we determined that long term storage (to date, up to 9 months) under these conditions allows unbiases RT-qPCR analyses. However, we note that repeated freeze and thaw cycles of stored specimens as well as purified RNA affect sample quality and result in gradually increasing C_T_ values.

### RT-qPCR

Quantitative analyses of SARS-CoV-2 (+)RNA from human specimen was carried out combining reverse transcription and qPCR in a one-step protocol using Luna Universal Probe One-Step RT-qPCR Kit w/o ROX (New England Biolabs, Cat. No. E3007E) on a Corbett Rotor-Gene 6000 instrument. Primers and probes are described above and listed (Table S1). Per reaction, each primer and probes were used at 500 nM. 2 µL of purified template RNA were used for each single reaction volume of 20 µL. RT-qPCR conditions were as follows: Reverse transcription (RT) (60°C/30 min), initial denaturation and Hot Start *Taq* polymerase activation 95°C/2 min, cycling (36x[denaturation 95°C/15 s, extension 60°C/30 s]) and final extension 68°C/5 min. 36 qPCR cycles were determined to largely avoid multiple unspecific byproducts and giving rise of sufficient amounts of biotinylated amplicon for subsequent pyrosequencing purposes. This amplicon could be used directly for pyrosequencing.

### Pyrosequencing

Pyrosequencing of the biotinylated single-stranded S-gene amplicon was performed on a PyroMark Q48 Autoprep device (Qiagen). The PyroMark Q48 Advanced CpG Reagents (4x 48) (Qiagen, Cat. No. 974002) including dNTPs, substrates and enzymes are loaded to the assigned cartridges with volumes being adapted to the assay requirements. The designed approach theoretically allows to read a SARS-CoV-2 (+)RNA-specific sequence fragment of 55 nt. Further, the sequencing primer (S_pbc_-CoV-2-S) was added to the assigned cartridge in a predetermined volume. 10 µL of each RT-qPCR amplicon were loaded to each well of a PyroMark Q48 Disc and then 3 µL PyroMark Q48 Magnetic Beads (300) (Qiagen, Cat. No. 974203) were added to each reaction. The sequencing reaction was initiated after denaturation and subsequent magnetic bead capture of the biotinylated single-stranded S-gene amplicon. The pyrosequencing software records the light signals corresponding to each well in the PyroMark Q48 Disc and saves the data graphically. Basecalled pyrograms were then manually inspected and analyzed for SARS-CoV-2 sequence verification.

## Data Availability

Not applicable

## List of abbreviations

(+)RNA: sense polarity [single-stranded] ribonucleic acid
CI: confidence interval
COVID-19: coronavirus disease 2019
MERS: Middle East respiratory syndrome
C_T_: cycle threshold
ORF: open reading frame
RdRP: RNA-dependent RNA polymerase
RT-qPCR: reverse transcription-quantitative polymerase chain reaction
SARS-CoV-1: SARS-CoV-2 (severe acute respiratory syndrome coronavirus 1
SARS-CoV-2: severe acute respiratory syndrome coronavirus 2
sgRNA: subgenomic RNA
*Taq*: *Thermus aquaticus*

## Declarations

### Ethics approval and consent to participate

For the collection and use of specimens at Helios University Hospital Wuppertal (North Rhine-Westphalia, Western Germany) and Klinikum Kassel (Hessen, Central Germany) with obtained approval of the Witten/Herdecke University Ethics board (covered by ‘CoronaKids’ [No. 61/2020] and No. 160/2020 for the use of routinely sampled and confirmed specimens for RT-qPCR/pyrosequencing method establishment). All work has been conducted according to the principles expressed in the Declaration of Helsinki.

### Consent for publication

Informed written consent was obtained from legal guardians of the involved children.

### Availability of data and material

Not applicable

### Competing interests

There are no conflicting interests for none of the authors, which need declaration.

### Funding

The study was financed by own institutional means.

### Authors’ contributions

PPW helped to design the methodology and helped to write the paper. PPW, JH and JP established RT-qPCR tests and pyrosequencing. FS with the help of JH conducted the tests on the pediatric cohort. AP supported RT-qPCR works and alternative sequencing methods. BG and JO provided specimens, technical support and helped in the study design and data interpretation. OM, AH and MA characterized the Wuppertal case of a Multisystem Inflammatory Syndrome in Children (MIS-C)/Kawasaki-like syndrome. ACR and DM supported sample preparation and RT-qPCR establishment. SW and MA designed, coordinated and contributed to data analyses with respect to the Wuppertal cohort study. ACWJ participated in the design of the pediatric cohort study, contributed specimens, helped in data interpretation, provided clinical case characterizations from Kassel and helped to write the paper. JP designed the RT-qPCR/pyrosequencing study concept, contributed to the pediatric cohort study, supervised the entire study, took part in protocol establishment, interpreted the data and wrote the paper.

## Acknowledgements

We thank Prof. Parviz Ahmad-Nejad (Institute of Medical Laboratory Diagnostics, Helios University hospital Wuppertal) and the involved staff, in particularly Jennifer Ortelt, Anna Wagener and Petra Menk, for providing specimens for method establishment and for the seamless collaboration.

## Supplemental information

### The supplemental information contains 1 Table and 4 Figures

**Table S1.**
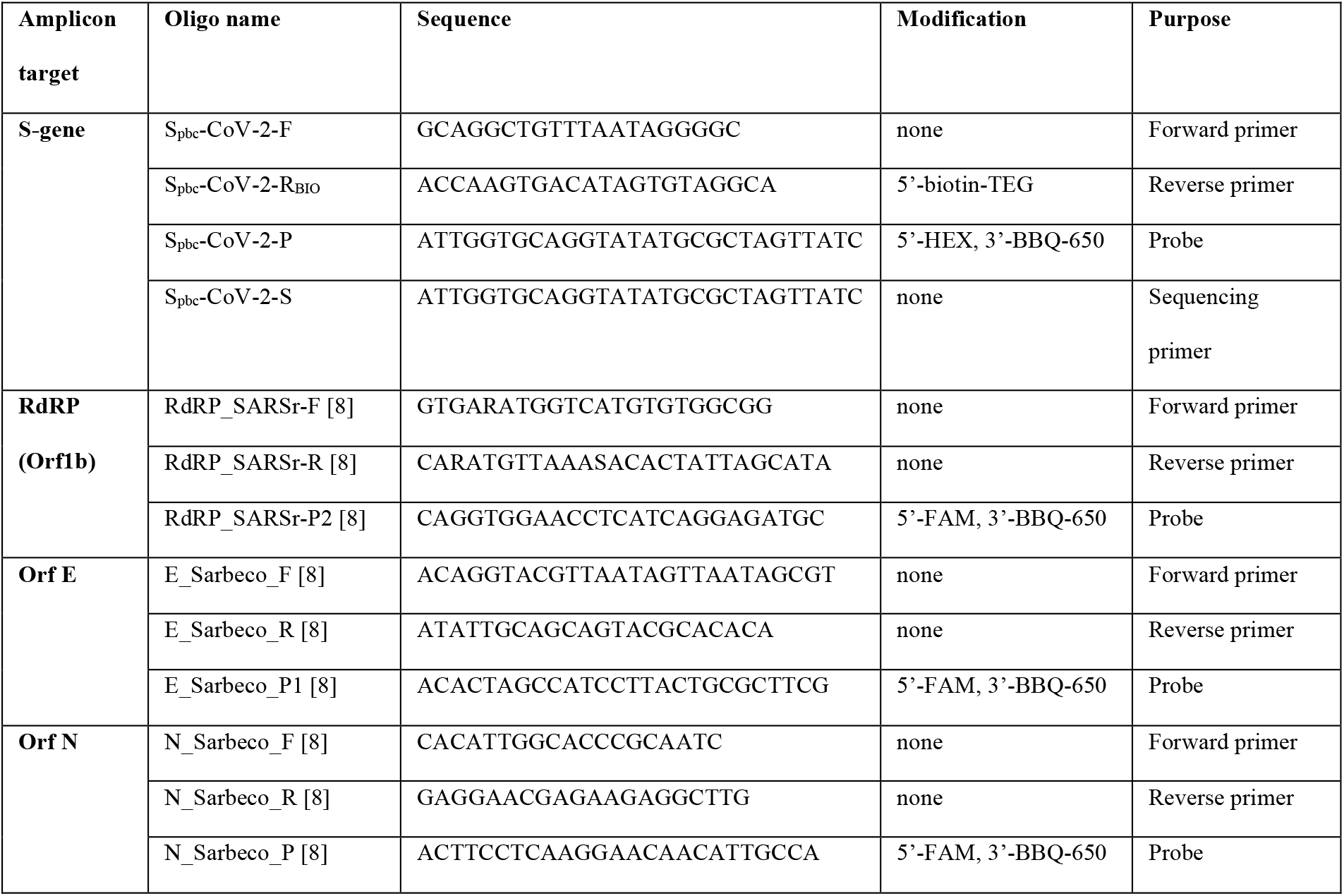
Oligonucleotides used for RT-qPCR and pyrosequencing.

**Figure S1.**
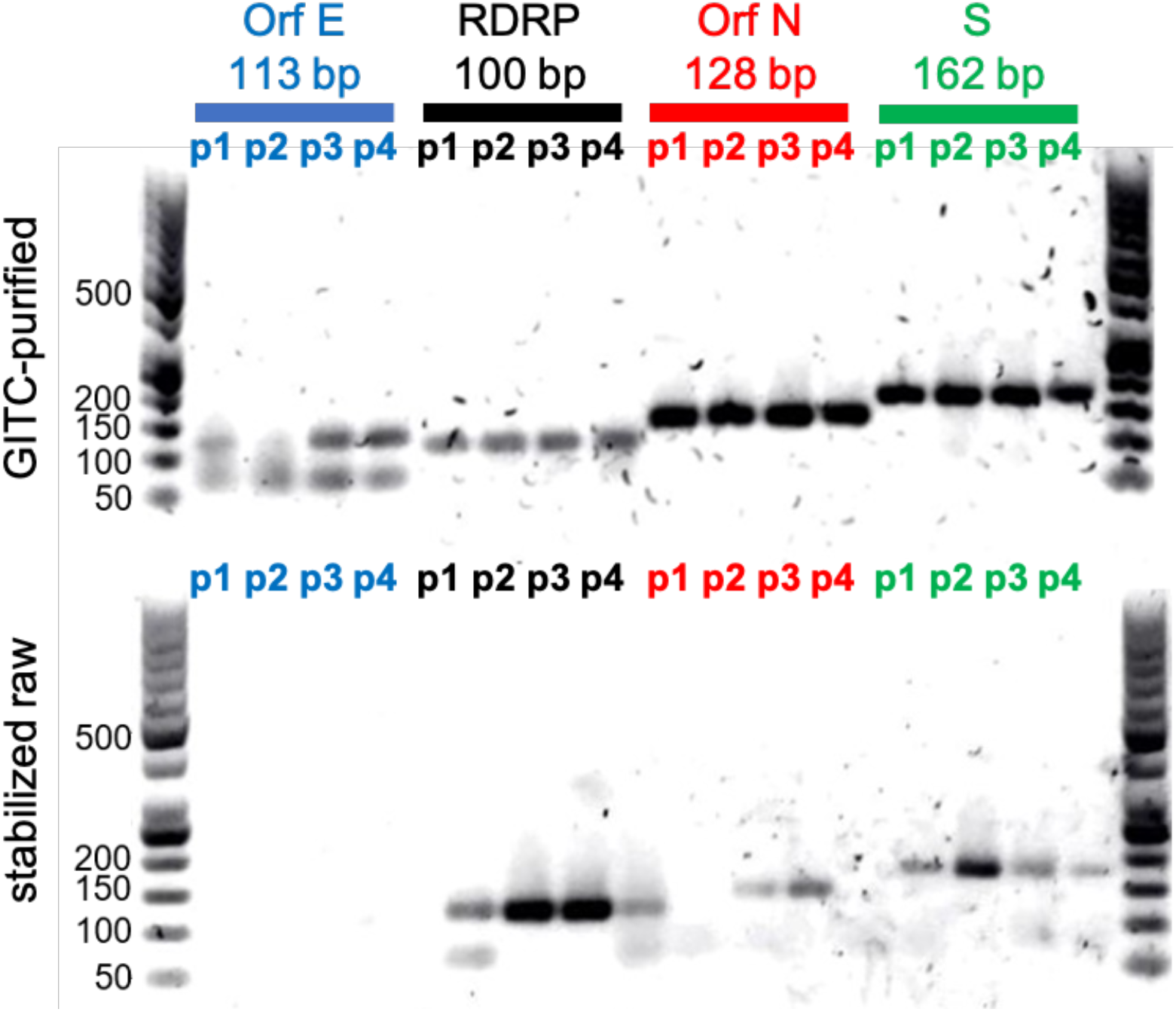
Results of semi-quantitative PCR for the performance comparison using GITC-purified RNA or stabilized raw specimens as RT-qPCR substrates. Four confirmed SARS-CoV-2-positive specimens (p1 to p4) were used in combination with primer pairs targeting Orf E, RdRP, Orf N and S-gene amplicons in singleplex qPCR reactions. From GITC-purified templates, amplicons with the correct size could be amplified from all specimens. Notably, we observed lower molecular weight byproducts for Orf E PCR. From stabilized raw specimens, in contrast, the Orf E amplicon was not amplified from none of the four samples. With varying band intensity, amplicons with correct sizes were amplified from stabilized raw samples for all cases for RdRP and S-gene targets and in some cases for the Orf N target. However, for unknown reasons, band intensities appeared less stable when compared with purified RNA samples. Moreover, we observed lower molecular weight byproducts in some cases.

**Figure S2.**
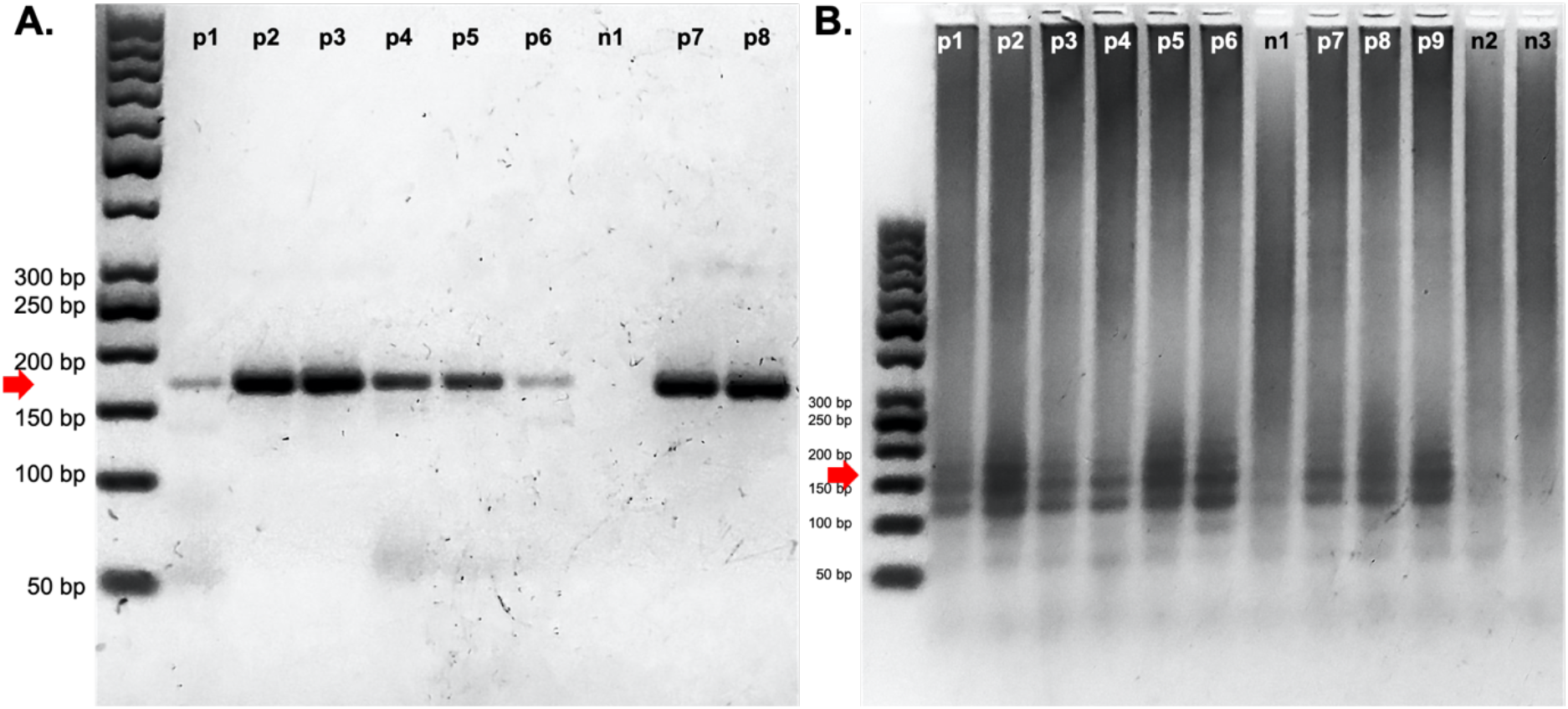
Results of semi-quantitative PCR of the SARS-CoV-2 S-gene amplicon. **A**. After 36 PCR cycles and subsequent agarose gel electrophoresis the specific 162 bp amplicon corresponding to the SARS-CoV-2 protein S-gene was visible (red arrow). The gel was loaded with 8 RT-qPCR samples from confirmed SARS-CoV-2-positive cases (p1 to p8) and 1 negative control (n1). The occasional weak appearance of PCR byproducts (see p1) seemed to correlate with relatively low viral (+)RNA load in the specimen. **B**. An excess of PCR cycles (40x) leads to enrichment of unspecific lower and higher molecular weight byproducts in all samples from confirmed positive samples (p1 to p9) as well as negative controls (n1 to n3). These byproducts severely impaired the successive pyrosequencing leading to ambiguous results.

**Figure S3.**
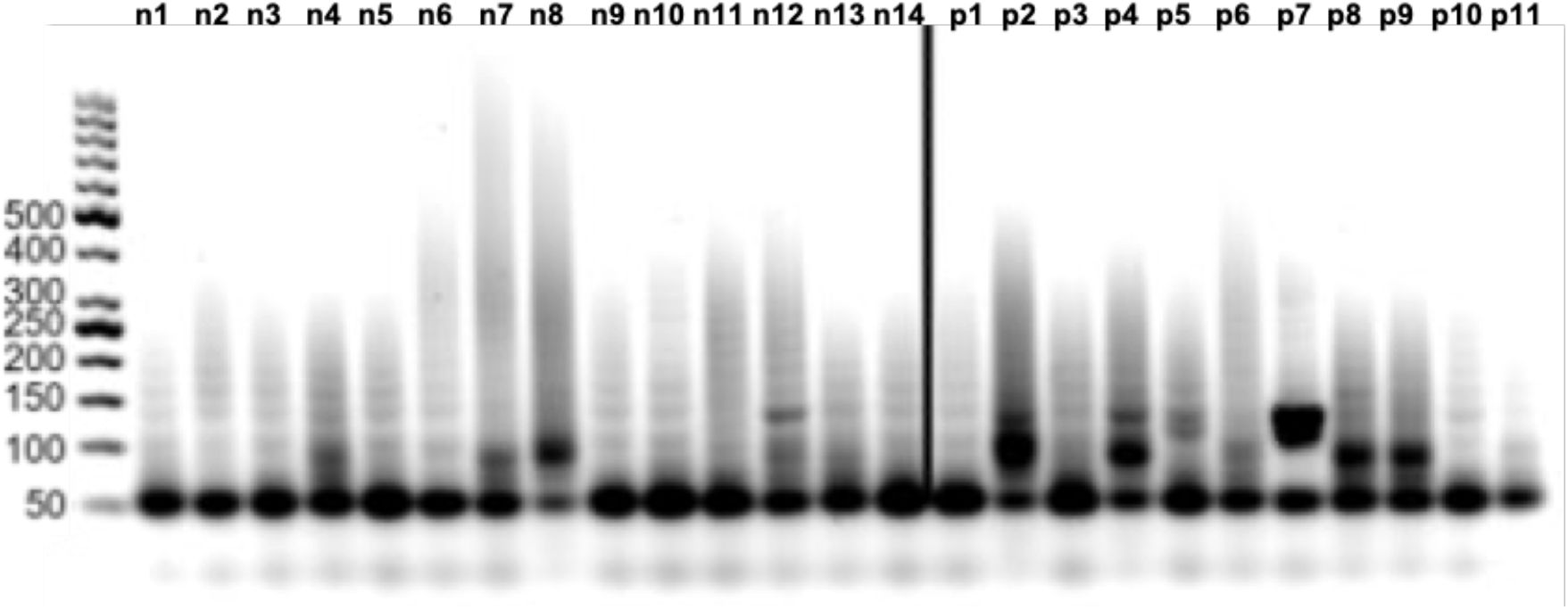
Results of a triplex PCR approach for the simultaneous detection of amplicon targets for RdRP, Orf E and Orf N. For comparison, 14 SARS-CoV-2-negative samples (n1 to n14) and 11 confirmed SARS-CoV-2-positive specimens were analysed by agarose gel electrophoresis. The simultaneous use of primers for RdRP, Orf E and Orf N amplicons leads to enrichment of unspecific lower and higher molecular weight byproducts in all samples, confirmed positives as well as negatives. Remarkably, in the fraction of negative samples bands of approx. 160 bp appear frequently.

**Figure S4.**
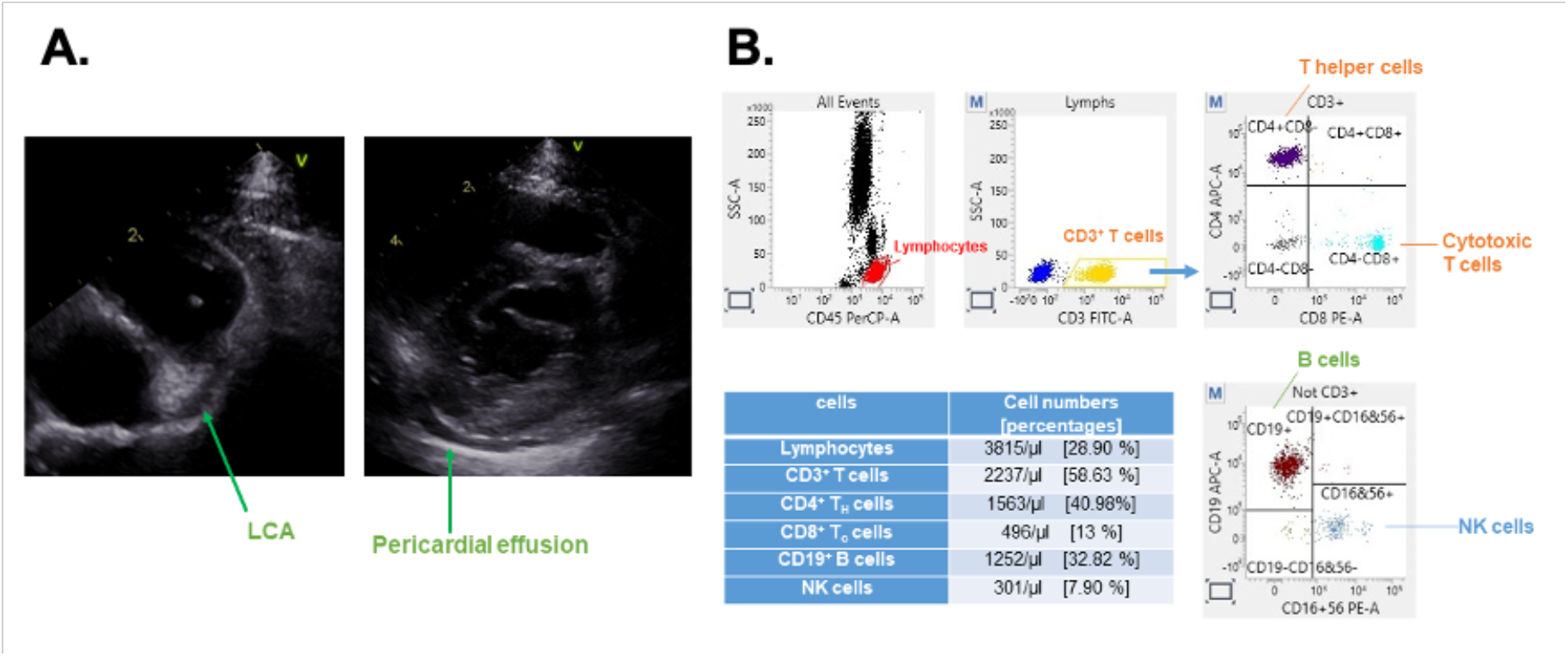
Presentation of a female toddler (age group 1-3 years) with a Multisystem Inflammatory Syndrome in Children (MIS-C)/Kawasaki-like syndrome associated with SARS-CoV-2 infection. **A**. Echocardiography revealed an enlargement of the left coronary artery (LCA) with a pericardial effusion. **B**. Flow cytometric characterization of the peripheral mononuclear blood cells resulted in normal ranges of CD3+ T, CD4+ T helper, CD19+ B, and CD16+CD56+ natural killer cells.

## Notes

### Competing Interest Statement

The authors have declared no competing interest.

### Author Declarations

For the collection and use of specimens at Helios University Hospital Wuppertal (North Rhine-Westphalia, Western Germany) and Klinikum Kassel (Hessen, Central Germany) with obtained approval of the Witten/Herdecke University Ethics board (covered by No. 61/2020 [CoronaKids] and No. 160/2020 for the use of routinely sampled and confirmed specimens for RT-qPCR/pyrosequencing method establishment). All work has been conducted according to the principles expressed in the Declaration of Helsinki.

